# Long non-coding RNAs (lncRNAs) NEAT1 and MALAT1 are differentially expressed in severe COVID-19 patients: An integrated single cell analysis

**DOI:** 10.1101/2021.03.26.21254445

**Authors:** Kai Huang, Catherine Wang, Christen Vagts, Vanitha Raguveer, Patricia W. Finn, David L. Perkins

## Abstract

Hyperactive and damaging inflammation is a hallmark of severe rather than mild COVID-19 syndrome. To uncover key inflammatory differentiators between severe and mild COVID-19 disease, we applied an unbiased single-cell transcriptomic analysis. We integrated a bronchoalveolar lavage (BAL) dataset with a peripheral blood mononuclear cell dataset (PBMC) and analyzed the combined cell population, focusing on genes associated with disease severity. Distinct cell populations were detected in both BAL and PBMC where the immunomodulatory long non-coding RNAs (lncRNAs) NEAT1 and MALAT1 were highly differentially expressed between mild and severe patients. The detection of other severity associated genes involved in cellular stress response and apoptosis regulation suggests that the pro-inflammatory functions of these lncRNAs may foster cell stress and damage. The lncRNAs NEAT1 andMALAT1 are potential components of immune dysregulation in COVID-19 that may provide targets for severity related diagnostic measures or therapy.

## Introduction

The Severe Acute Respiratory Syndrome Coronavirus 2 (SARS-CoV-2) pandemic continues around the world,(1,2) but the underlying pathophysiology of coronavirus disease 2019 (COVID-19) is ill-defined. Symptoms and progression of COVID-19 vary widely(3) as some patients may be asymptomatic while others exhibit disease with varying severity.(4,5) Common symptoms include fever, cough, and fatigue, which generally appear 2 to 14 days after exposure,(6,7) while rarer symptoms include dyspnea, headache/dizziness, nausea, diarrhea, and hemoptysis.(8) Severe cases of COVID-19 are distinguished by strong inflammatory responses that can lead to multiorgan damage and death.(9) The mechanisms that separate mild and severe disease remain poorly understood.

After viral exposure, the inflammatory response to COVID-19 commences with signaling cascades that lead to secretion of type I interferons, cytokines and chemokines.(10) This initial exposure also activates inflammasomes, multimeric protein complexes that play an important role in triggering inflammation and the subsequent initiation of an adaptive immune response.(11–13) The Nod-like receptor pyrin domain-containing 3 (NLRP3) inflammasome is a major cause of cytokine storm associated the with clinical manifestations of severe COVID-19 disease.(14) Furthermore, coronavirus viroporin proteins activate the NLRP3 inflammasome which regulates the secretion of IL-1β and IL-18.(15) Pyroptosis, a programmed cell death pathway that leads to immune cell depletion, is also regulated by activation of the NLRP3 inflammasome and is an important mechanism of viral pathogenesis in both SARS-CoV-2 and SARS-CoV.(16–18) These studies suggest that investigation of inflammasome regulation may elucidate understanding of COVID-19 disease pathophysiology.

Single-cell studies of COVID-19 patients have found dysregulated immune compartments in the respiratory tract as well as peripheral blood.(19–24) However, it is challenging to directly compare results across studies in different tissues due to differences in cell cluster identification between physiological compartments. We postulated that simultaneous analysis of severe versus mild COVID-19 patients across respiratory and peripheral immune compartments using integrated clustering would uncover overall effectors of immune dysregulation in the COVID-19 immune response. To achieve this goal, we integrated single-cell datasets, one from bronchoalveolar lavage (BAL) and one from peripheral blood mononuclear cells (PBMC),(19,20) in order to examine disease transcriptomics across severities as well as between local and peripheral cellular environments. We utilized an unbiased analytical strategy that was agnostic to specific gene functions and focused on genes with severity dependent expression across different cell types. Taken together, we uncovered genes contributing to the dysregulated COVID-19 immune response prominent in severe relative to mild disease. Moreover, we identified cell types where these inflammatory regulators manifest in local and peripheral compartments.

## Methods

### Dataset preprocessing and integration

We selected publicly available single-cell datasets with patient severity metrics and ample sequencing depth to pass our quality filters for integration into our combined dataset. The raw count matrices for BAL cells and PBMC cells were downloaded from the NCBI Gene Expression Omnibus (accession number GSE145926) and the COVID-19 Cell Atlas (https:/www.covid19cellatlas.org/#wilk20), respectively. Patients who were mechanically ventilated or had PaO_2_/FiO_2_ ⍰≤⍰300 mmHg indicating hypoxemia consistent with acute respiratory distress syndrome (ARDS)(25) were designated as severe patients while all others were considered to have mild disease **(Table S1)**. The BAL dataset contained three healthy controls, while the PBMC dataset contained six **(Table S2)**. Both datasets were preprocessed using the R program Seurat.(26) Briefly, cells were filtered to only include cells with unique molecular identifier (UMI) counts greater than 1000, gene count between 200 and 6000, and less than 10% of genes mapping to mitochondrial genes. The function SCTransform from the Seurat package was applied to each dataset separately to regress out technical variability as well as the percentage of mitochondrial gene expression.(27) Transformed BAL and PBMC datasets were integrated with 3000 integration features and 50 integration anchors as recommended in Seurat.(28) We found that the “M3” mild patient sample from the BAL dataset contained only 369 total cells, while every other patient sample for BAL or PBMC had at least 1200 cells. The M3 sample was removed before differential expression analysis to avoid skewing results due to extremely low cell counts.

### Clustering and Identification

The integrated dataset was dimension reduced using principal component analysis (PCA) and clustered with a resolution set to 0.5 and including the top 30 principle components. The clustering was visualized using uniform manifold approximation and projection (UMAP).(29) The raw integrated dataset was normalized by applying SCTransform to the full integrated dataset. This normalized count matrix is utilized for all subsequent analysis. Marker genes for each cluster were computed using the FindAllMarkers function with the Model-based Analysis of Single-cell Transcriptomics algorithm (MAST) for differential expression with UMI count as a latent variable.(30,31) Cluster markers were then inspected and labeled according to known cell markers(Supplemental file 1).(32,33) A second round of clustering with a resolution of 1 was then conducted to further classify subtypes of identified cells. Clusters with fewer than 300 cells were reassigned to larger clusters using Seurat integration label transfer. Cluster identities were scored and verified using a signature matrix generated from flow cytometry sorted RNA-seq data of immune cells.(34) Plots of cell clusters and key cell type markers were generated using Seurat’s plotting functions.

### Cell Proportions

Cell types were tallied for each sample, and the percentage abundance of each cell type was calculated. Cell proportions for healthy controls, mild patients, and severe patients were compared using a two-sided pairwise test of equal proportion with false discovery rate (FDR) p-value adjustment. The resulting proportions were plotted using the ggplot2 R package.(35)

### Differential Expression

For each cell type, differentially expressed genes (DEGs) were calculated separately for BAL and PBMC cells using MAST with UMI count as a latent variable. To support MAST differential gene expression analysis between three sample groups, the Seurat built-in differential expression function FindMarkers was modified to generating iterations of the hurdle model corresponding to each set of two compared conditions. Genes from each cell type that were differentially expressed across all three comparisons between healthy controls, mild, and severe patients were tallied. For BAL, only genes with FDR adjusted p-values less than 1e-7 across all comparisons were considered (17.4% of all DEGs in BAL), while PBMC DEGs were considered if all FDR adjusted p-values across comparisons for that gene were less than 0.05 (16.4% of all DEGs in PBMCs). The difference in p-value threshold was used to filter a similar proportion of genes from BAL and PBMC data due to the smaller number of DEGs detected in PBMCs (200-400 in PBMCs, 1000+ in BAL). These highly differentially expressed genes were further filtered by removing genes that were not differentially expressed across all conditions in at least 5 different cell types (1/3 of our total cell types). The lists of PBMC and BAL highly differentially expressed genes were then combined, removing duplicates.

Some of the genes identified were highly cell type specific. These genes also had the highest residual variance. Since these genes represent intrinsic cell type differences rather than biologically interesting differentially expressed genes, they were removed. To determine this filter threshold, the residual variance of the top 100 variable genes were plotted in decreasing order to determine the “elbow” point where the variance stops decreasing at a rapid rate.**(Figure S1)** This resulted in the 21 most variable genes being removed. We termed the 50 remaining differentially expressed genes recurrent differentially expressed genes (rDEGs) since they were found in multiple cell types and showed differential expression between patients and healthy controls as well as between severities. The rDEG expression data was exported to Monocle 3 to generate modules for gene ontology (GO) enrichment analysis. We generated 4 modules from the 50 rDEGs using the find_gene_modules function in Monocle 3 with 30 principle components and a resolution of 0.8.(36–38) Differential module expression was calculated using ANOVA using aggregated module expression levels and processed with a tukey posthoc test. Module gene ontology enrichment was computed topGO with default settings.(39) Module and gene level plots were generated using the R packages ggplot2, ComplexHeatmap, and Circlize.(35,40,41)

### Validation

We compared rDEG trends from our analysis with two additional COVID-19 datasets, one with nasopharyngeal data to compare against the local inflammatory environment of our BAL data, and one in PBMCs to compare against the peripheral environment in our PBMC data.(22,23) Cell types from our analysis were transferred onto the validation datasets using Seurat to identify the validation cells for comparison. Each validation dataset was filtered and preprocessed separately using the same parameters as the main dataset. After preprocessing and label transfer, DEGs were generated independently for each validation set for our transferred cell types. We compared the rDEGs we focused on in the manuscript with DEG results from each validation set, noting whether the same DEG was detected and whether the direction of change was the same.

## Results

### Integrated PBMC and BAL analysis identified 26 clusters consolidated into 15 cell types

After quality filtering, we recovered 100,739 single cell transcriptomes. From these, we recovered 26 cell clusters from Seurat. **(Figure S2)**. Since we identified cell types from the integrated dataset containing both PBMC and BAL cells, we were able to examine how each of our cell clusters behaves across both physiological compartments. The clusters did not aggregate based on sample type or patient condition **(Figure S2)**, indicating successful integration clustering of the two datasets.

From the 26 clusters, 11 were identified as monocyte/macrophage (Mo/Ma) clusters. Since our clusters contain both monocytes from the PBMC sample as well as macrophages from the BAL, we designated them as MoMa clusters. Six of the MoMa clusters showed classically M1 associated transcriptomes with increased expression of VCAN, FCN1 and CD14 expression.(42,43) These clusters also expressed other pro-inflammatory factors such as S100A8, CCL2, CCL3, CCL7, and CCL8.(44,45) Three other MoMa clusters showed M2 polarization with increased FN1 expression along with decreased VCAN and FCN1 expression. These M2 MoMa clusters also expressed Th2 associated inflammatory factors such as MRC1 and CCL18.(45) All MoMa clusters expressed FCGR3A (CD16a).(42) Two additional clusters were labeled as intermediate MoMa because they did not show distinct transcriptomes corresponding to either M1 or M2 groups. One intermediate MoMa cluster overexpressed MALAT1, while the other overexpressed metallothionein proteins including MT1F and MT1G.

We also identified two clusters of CD4+ T cells (CD4 and IL7R), one cluster of T regulatory cells (IL2RA and LAG3),(46,47) and three clusters of CD8+ T cells (CD8A). Two of the CD8+ T cell clusters were labeled as CD8+ memory cells due to their high CCL5 and GZMH expression.(48) Other identified immune cell clusters include natural killer (NK) cells (SPON2 and NCAM1), neutrophils (NAMPT),(34) naïve B cells (MS4A1), plasmablasts (IGJ and MZB1), plasmacytoid dendritic cells (IRF8 and PLD4),(49,50) and myeloid dendritic cells (CD1C)and LGALS2).(34,51) In addition to immune cell types, we also found two epithelial clusters. One contained a mixture of epithelial and granulocyte markers including KRT19 and SLPI(34,52) while the other also contained the additional markers PPIL6 and CFAP300 for pneumocytes and ciliary cells, respectively.(53,54)

The 26 clusters were consolidated into 15 cell types (**Figure S2)** to streamline further analysis by combining clusters that are not distinguishable when examining their canonical marker expression levels. This consolidation also prevents cell groups with many clusters such as the M1 MoMa group from overshadowing those with fewer clusters in our subsequent differential gene expression ranking analysis. Cells within each cluster were compared against their original identifications from both their respective dataset, and most clusters identifications were consistent. The one exception was the intermediate MoMa type, which was predominantly composed of macrophages from BAL, but also contained a mixture of monocytes, CD4+ and CD8+ T cell identifications from PBMCs **(Table S3)**.

### Proinflammatory cell types are enriched in severe COVID-19 patients

Cell type proportions within BAL and PBMC sample groups showed distinct differences between patients and healthy controls as well as between mild and severe patients **(Figure 1)**. In BAL samples of patients with severe disease, proinflammatory cells such as M1 MoMa and neutrophils showed increased abundance (p<<<.01, **Table S4**), while immunoregulatory cell types including M2, intermediate MoMa, and Tregs were less abundant (p<<<.01). BAL samples of patients with mild disease showed decreased abundance of M1 MoMa and neutrophils and increased Tregs and CD8+ memory T cells compared to healthy controls and severe patients.

**Fig 1:**
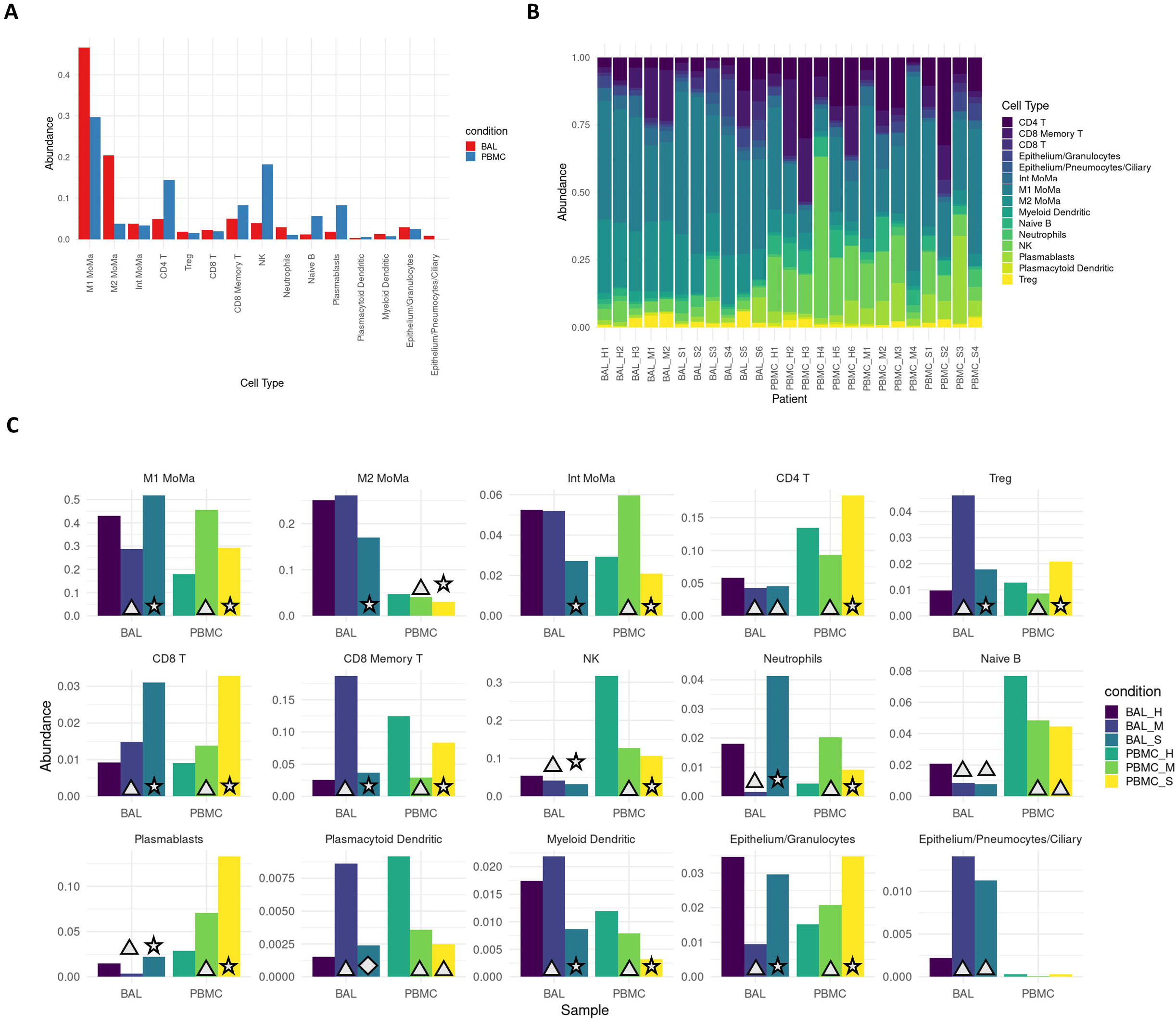
Severe patients show increased proportions of proinflammatory cell types. **A**. Overall average abundance of each major cell type for all cells. **B**. Per patient abundance of all major cell types for all cells. **C**. Per cohort (bronchoaveolar lavage (BAL) and peripheral blood mononuclear cells (PBMC)) and per condition (healthy, mild, severe) abundance for each cell type. Conditions that are significant versus their respective controls are labeled with a triangle (p<.05). Conditions that are significant between severe and both mild as well as healthy controls are labeled with a star (p<.05). Conditions that are significant between severe and mild, but not between severe and healthy controls are labeled with a diamond (p<.05).

In PBMCs, the trends in M1 MoMa and neutrophils are reversed. Tregs and CD8+ memory T cells are less abundant in PBMCs of mild patients. These opposing patterns may illustrate heavy recruitment of the cell types abundant in BAL, resulting in depletion in the PBMCs that results in an increase in relative abundance of non-recruited cells in PBMCs. Mild patients also showed an increase of intermediate MoMa in PBMCs, reinforcing the pattern of relative increases in abundance of immunoregulatory cell types in mild patients in both BAL and PBMC compartments.

### Recurrent DEG (rDEG) modules highlight key pathways in COVID-19 immune response

We identified an average of 1158 DEGs per cell type for BAL samples, and 260 DEGs per cell type for PBMC samples.**(Table S5**,**S6)** After filtering, we identified 50 rDEGs across our 15 cell types that formed 4 distinct modules **(Figure 2)**. Module 1 showed significant GO enrichment for developmental processes (p<.05) but did not show differential expression between conditions. Module 2 showed significance for viral defense and Type I interferon GO terms. Most genes in this module were interferon induced genes including the first three IFIT family genes, ISG15, CXCL10, and MX1. This module was significantly overexpressed in BAL of all patients versus healthy controls (p<.01).

**Fig 2:**
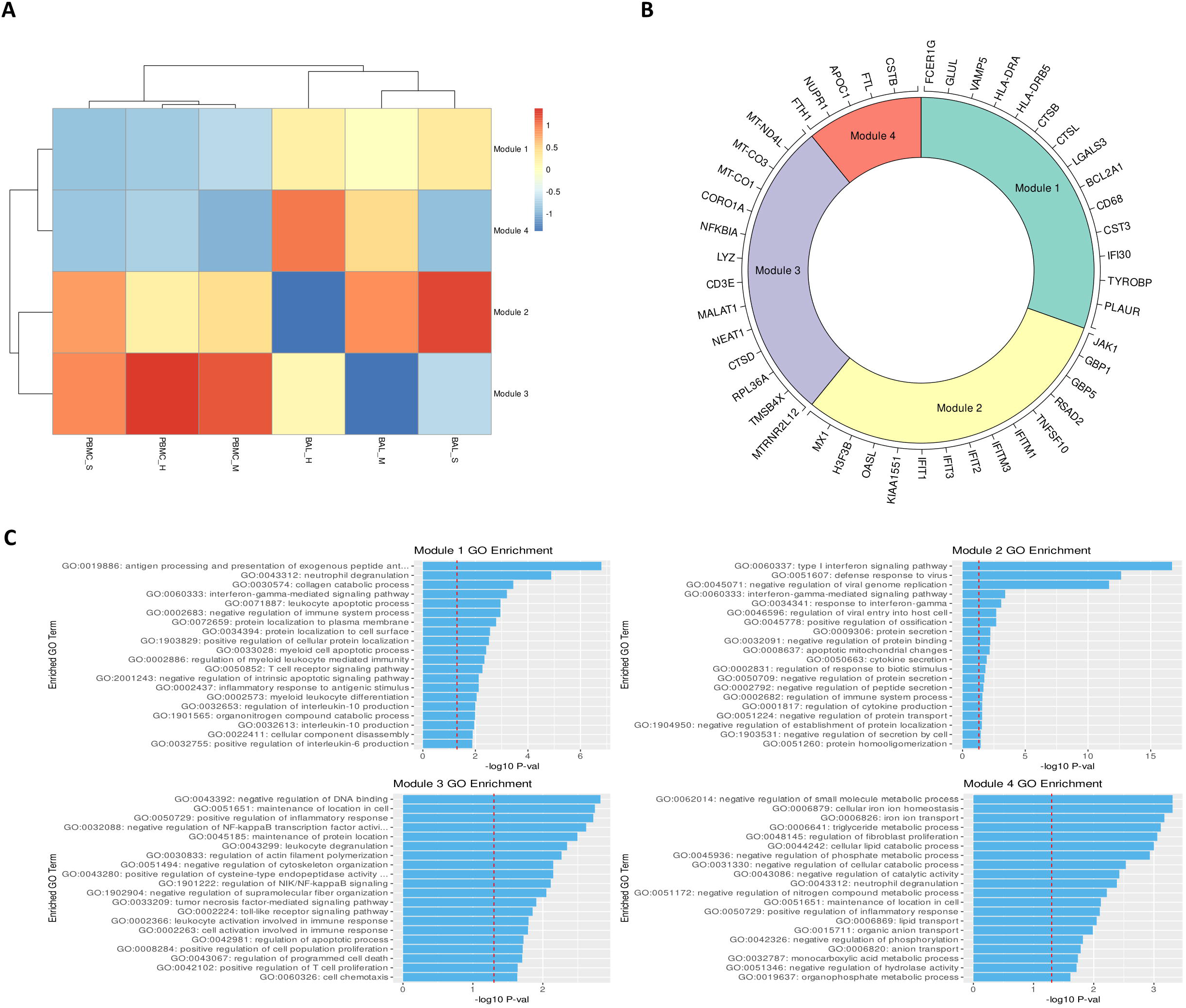
rDEGs grouped into four distinct modules with immune regulation enriched GO terms. **A**. Heatmap of modules generated from the recurrent differentially expressed geens (rDEGs). Triangles indicate significance (p<.05) versus healthy control within the sample cohort; diamonds indicate significance between severe and mild patients (p<.05). **B**. Module membership for each module. Modules 1 and 4 contain a mixture of metabolic and immune response related genes. Module 2 contains genes related to interferon activated viral defense. Module 3 contains other inflammatory regulation genes and stress response genes (generated using the Circlize R package). **C**. Per module GO term enrichment showing the top enriched terms for each module and their respective p-values with the red line indicating -log10(0.05). The first three modules contain inflammation related terms in their most enriched terms, while module 4 only contains metabolism related terms.

Module 3 was enriched for macromolecule synthesis and cellular processes. This module includes the immunomodulatory lncRNAs NEAT1 and MALAT1.(55,56) It also includes MTRNR2L12, an anti-apoptotic lncRNA, and NFKBIA which is an NF-κB inhibitor. The module was significantly underexpressed in BAL of mild patients versus healthy controls, and it was overexpressed in BAL of severe patients versus mild patients. Module 4 had significant terms related to negative regulation of metabolic processes. This module included the NUPR1 stress response gene, and CSTB which is an inhibitor of cathepsins like CTSL and CTSB that are involved in COVID-19 viral entry.(57) Module 4 was significantly underexpressed in BAL of severe patients versus healthy controls.

### Stress response, apoptosis, and viral entry related genes show severity dependent expression

We analyzed individual rDEGs in each of our five most abundant cell types: M1 MoMa, M2 MoMa, CD4+ T cells, NK cells, and CD8+ memory T cells (**Figure 3**). This analysis confirmed previous reports of downregulation of HLA genes(20,58) such as HLA-DRA and HLA-DRB5 in COVID-19 patients, with severe patients showing the most downregulation. We also saw upregulation of interferon related genes including MX1, and IFIT1-3. This increase was greatest in mild patients, correlating with previous findings of immune exhaustion (**Table S7**).(59,60) Further examination showed additional severity dependent patterns of differential expression of transcripts related to the stress response, cell death, and viral entry in cell types involved in the viral immune response.

**Fig 3:**
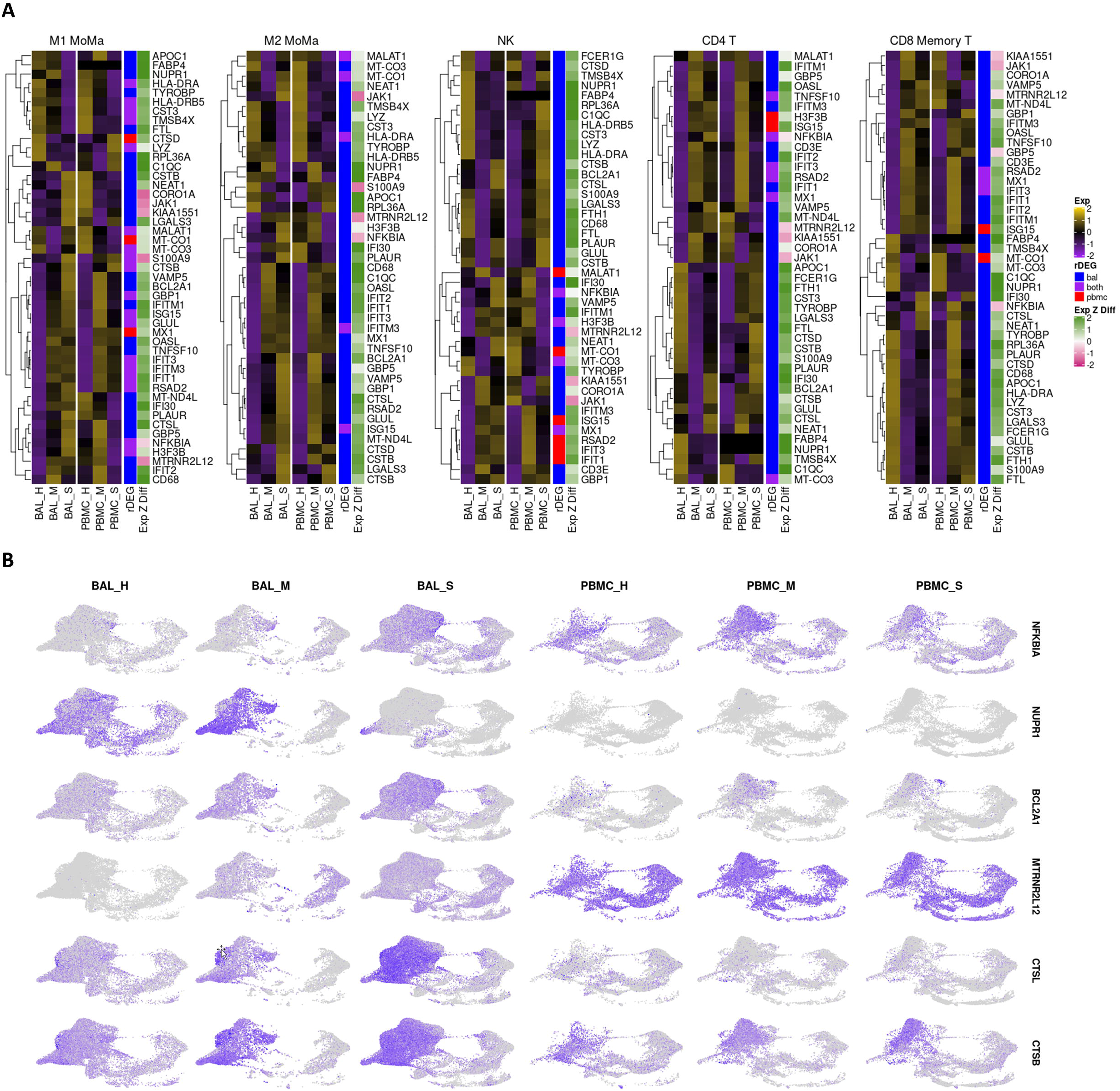
rDEG expression in most abundant cell types highlights differential immune regulation between mild and severe patients in both BAL and PBMC cohorts. **A**. Heatmaps visualizing rDEGs within each of the top five most abundant cell types in our dataset (generated using the ComplexHeatmap R package). For each cell type, the full rDEG list was filtered via the same p-values (p<10e-7 for BAL, p<.05 for PBMC) and only rDEGs that are differentially expressed below these thresholds for either BAL or PBMC are included in the plot. Expression levels are normalized separately for each cohort. The first sidebar indicates which cohort the particular gene passed the rDEG threshold for, while the second sidebar indicates the ratio of expression of the particular gene between BAL and PBMC with green (positive values) indicating higher expression levels detected in BAL. **B**. Visualization of select rDEGs representing pathways outside of the main interferon activated gene group that are relevant to disease. These genes are visualized separately for each cohort and condition using the sample UMAP projection of cell types from Figure 1. Each gene shows cell type, cohort, and condition specific differences in localization across the dataset.

The NF-κB inhibitor NFKBIA was upregulated in all five most abundant cell types within the BAL of severe patients compared to healthy controls and mild groups. In PBMCs of severe patients, NFKBIA was downregulated compared to healthy controls and mild patients except in CD8+ memory T cells. This pattern of localized overexpression in BAL may indicate increased NFKBIA activity in response to local hyperactivity of NF-κB. Furthermore, the stress response gene NUPR1, whose downregulation leads to cell death, was downregulated in M1 and M2 MoMa in the BAL of severe patients and upregulated in mild patients, indicating a pro-apoptotic shift in severe patient MoMa clusters. NUPR1 was downregulated in BAL of both mild and severe patients for NK cells, CD4+ T cells, and CD8+ Memory T cells.

Mild and severe patients also had variable expression of two anti-apoptotic genes, the BCL2 inhibitor BCL2A1 and the lncRNA MTRNR2L12. BCL2A1 was significantly upregulated in BAL of severe patients over healthy controls and mild groups for M1 and M2 MoMa, NK cells, and CD4+ T cells. Mild patients showed downregulation of BCL2A1 versus healthy controls in NK and CD4+ T cells. Additionally, MTRNR2L12 was upregulated in BAL of both mild and severe patients in M1 and M2 MoMa, NK cells, CD4+ T cells and CD8+ Memory T cells. The upregulation of these anti-apoptotic genes shows a defensive response to apoptotic cell stresses, particularly in BAL.

CTSL, which is a critical protein in the viral entry pathway for COVID-19, was upregulated in BAL of severe patients in M1 and M2 MoMa in mild patients and healthy controls. This suggests a faster viral entry pathway in severe patients, which may contribute to the formation of a hyperinflammatory response. In BAL of NK, CD4+ T cells, and CD8+ Memory T cells, CTSL was downregulated in mild patients and upregulated in severe patients. CTSB, also implicated in viral entry, showed similar patterns.

### NEAT1 and MALAT1 are differential regulators of inflammation in severe COVID-19

The pro-inflammatory lncRNA NEAT1 passed our rDEG threshold in BAL samples for nine different cell types, more than any other gene in our analysis. These cell types include M1, M2 and intermediate MoMa, NK cells, CD4+ T cells, CD8+ memory T cells, naïve B cells, myeloid dendritic cells, and epithelium/basal cells (**Figure 4**). NEAT1 is localized to the site of infection and inflammation since it is not differentially expressed in PBMCs. Additionally, among rDEGs, it has one of the highest averages in log2-fold change between severe and mild patients (**Figure 4**). NEAT1 is overexpressed in BAL of severe patients and underexpressed in mild patients. The epithelial/basal cell group is the exception where mild groups also show NEAT1 overexpression over healthy controls, but expression is still significantly higher in severe patients versus mild patients.

**Fig 4:**
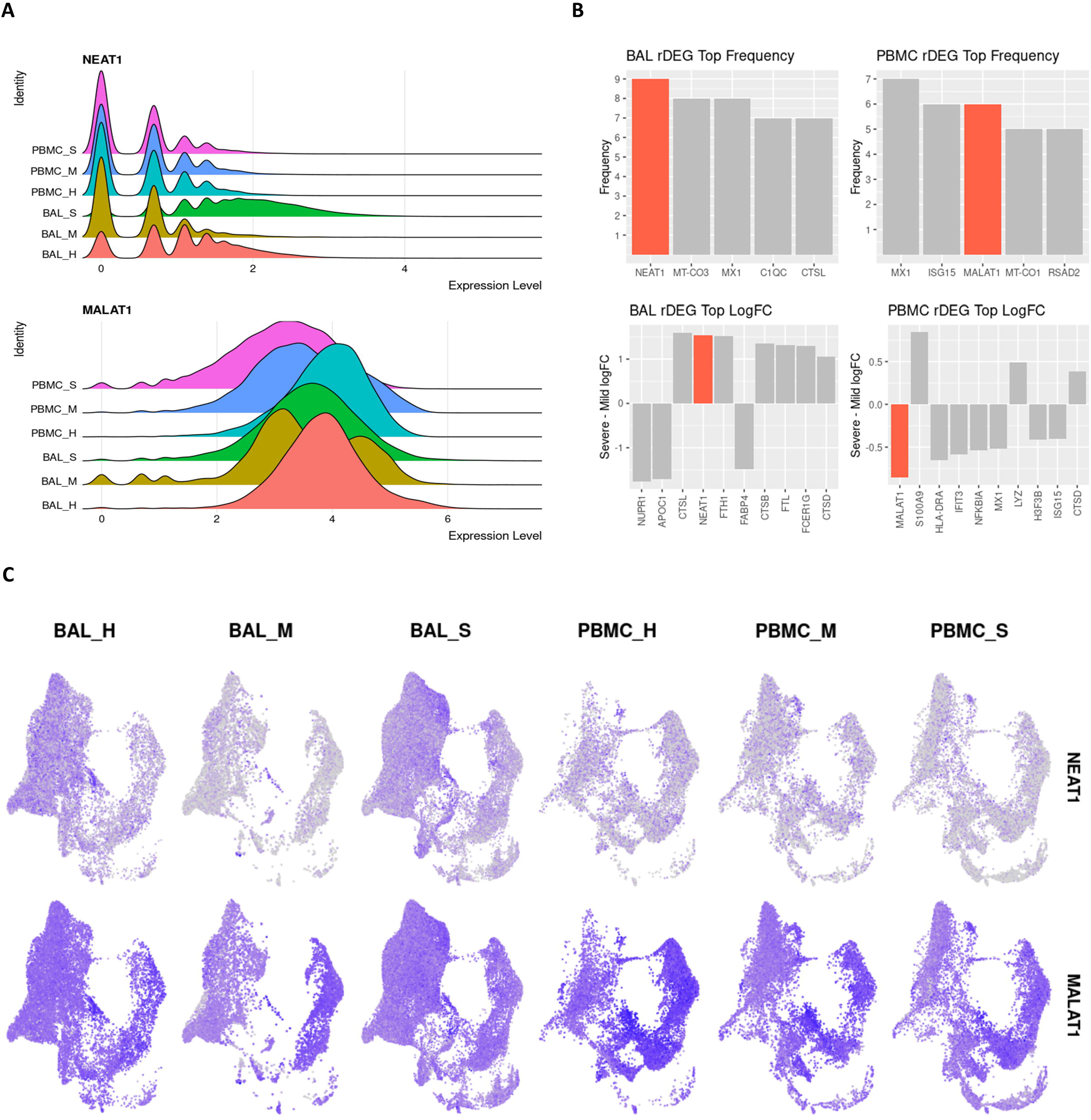
lncRNAs NEAT1 and MALAT1 are strongly differentially expressed between severe and mild patients and represent key inflammatory regulators in BAL and PBMC respectively. **A**. Violin plots showing overall expression level density across patient conditions in the entire dataset. Even at the full dataset scale, these distributions show that NEAT1 is overexpressed in BAL of severe patients while MALAT1 is underexpressed in PBMCs of severe patients. **B**. Frequency of detection across cell types for rDEGs shows NEAT1 as the most detected rDEG in BAL, with MALAT1 tied for second among rDEGs in PBMC. The top log2-fold change of rDEGs in severe versus mild patients also shows NEAT1 and MALAT1 among the rDEGs with the highest absolute change between severe and mild conditions. **C**. Visualization of NEAT1 and MALAT1 via UMAP projection shows more cell type localized expression in NEAT1. It is also clearly underexpressed in mild BAL cases. MALAT1 also shows a more subtle but significant underexpression in severe patient PBMCs.

Another immunomodulatory lncRNA, MALAT1, was the second most frequent rDEG in PBMCs. It passed our rDEG threshold in 6 cell types (tied with ISG15) and 3 cell types in BAL. In BAL derived M1 and M2 MoMa, MALAT1 was underexpressed in mild patients compared to both healthy controls and severe patients. In CD4+ T cells, MALAT1 shows consistent overexpression in mild patients and underexpression in severe patients. In PBMCs, MALAT1 was underexpressed in severe patients versus both healthy controls and mild patients in M1, M2 and intermediate MoMa, NK cells, plasmablasts, and epithelial/basal cells.

### Validation of rDEG expression patterns

We projected the cell type classifications in our analysis via Seurat’s label transfer feature to two other COVID-19 datasets, one with nasopharyngeal samples(22) and one with PBMC samples(23). Comparison of cell labels from the original datasets versus our transferred labels shows general agreement among the cell IDs, allowing us to use our cell type labels for direct comparison of rDEG patterns in the validation data **(Figure S3&S4)**. We compared rDEG expression patterns of the genes analyzed in the previous two sections in the same five cell types. Among each gene’s statistically significant changes in our analysis between healthy, mild, and severe cases, 36.3% were significant in our validation cohorts. However, more than two thirds of the non-significant findings in validation were from comparisons with healthy control in the nasopharyngeal dataset. This is likely due to the small number of cells recovered from these controls after filtering. Only 1148 cells were recovered from control samples after filtering compared to 35715 and 25546 for moderate and severe cases respectively. Among the findings that were significant in both our data and in validation, we found that 79% were significant in the same direction. Notably, NEAT1 showed 100% agreement in validation of BAL, while MALAT1 showed 100% agreement in validation of PBMCs **(Table S8**,**S9 & S10)**.

## Discussion

Our analysis of BAL and PBMC single cell data in COVID-19 patients has elucidated key differences between mild and severe disease. We were able to combine cells from both PBMC and BAL in an integrated analysis. Although our intermediate MoMa group had a mixed group of PBMC cells, our overall identifications were consistent across both datasets. Furthermore, the cells in the intermediate MoMa group consisted of cells with weak expression of a wide range of canonical markers. These cells may be intermediate immune cells from different lineages that share a similar transcriptomic profile. By conducting analysis simultaneously on cells from the local infection site in the lung as well as the peripheral immune system, we contrast how the disease manifests and interacts across both compartments. We have identified differentially expressed genes that vary with severity, are highly differentially expressed across multiple cell types, and represent key functions related to the hyperinflammatory disease state. NEAT1 was the most widely differentially expressed gene across cell types within BAL; it also exhibited a high log-fold change that correlated with disease severity. The ubiquity of NEAT1, its specific localization to BAL cells, and pro-inflammatory functions suggests that it may be a key mediator of the inflammation seen in severe COVID-19. NEAT1 is a well characterized activator of the NLRP3 inflammasome, as well as NLRC4 and AIM2 inflammasomes, which in turn amplify the inflammatory response.(55) However, an overactive immune response contributes to lasting tissue damage in severe COVID-19 disease. Intense inflammation through activation of the NLRP3 inflammasome can also lead to pyroptosis, driven by the upregulation of NEAT1.(55,61) These highly inflammatory and damaging effects of NEAT1 illustrate how overexpression in severe patients might lead to the inflammatory tissue damage seen in severe COVID-19.

MALAT1 also exerts various immunological effects including the mediation of NLRP3 inflammasome activation.(62,63) MALAT1 has been linked to M1-like activity in macrophages, promoting inflammation.(64) Our finding that MALAT1 is overexpressed in BAL MoMa of severe versus mild patients suggests that it might be involved in precipitating a shift towards M1 macrophages that exacerbates inflammation. This is further supported by our findings that severe patients show expansion of M1 macrophages and decrease of M2 and intermediate macrophages in BAL, while mild patients show decrease of M1 macrophages. Furthermore, MALAT1 was overexpressed in CD4+ T cells of mild patients. This is also reflective of MALAT1’s protective role in T cells. Loss of MALAT1 expression has been shown to push T cells towards the inflammatory Th1 and Th17 phenotype while also decreasing Treg differentiation.(65) This function matches our observed increase in abundance of Tregs in mild patients. Thus, the upregulation of MALAT1 we see in mild patients may be contributing to the more subdued immune response observed in these patients.

The severity dependent differential expression of other genes in our analysis provides further evidence of increased cellular stress reflective of a NEAT1 and MALAT1 enhanced hyperinflammatory state. NF-κB is induced in COVID-19 infection.(66) Although we did not detect differential activity of NF-κB directly, we found upregulation of its inhibitor NFKBIA in BAL of severe patients which suggests a feedback response to strong NF-κB activity. NFKBIA’s downregulation in PBMCs of severe patients may be due to localization of cells expressing NFKBIA to the site of infection in attempts to regulate the hyperactive inflammatory state.(67) The upregulation of BCL2A1 and MTRNR2L12 is also indicative of extensive cellular stress.(68,69) While MTRNR2L12 is upregulated in both mild and severe disease, BCL2A1 is upregulated exclusively in severe disease. The increased activity of these anti-apoptotic genes, particularly in BAL of severe patients, shows additional evidence of the cellular stress induced by infection and inflammation. These genes may be responding to pyroptosis pathways triggered by inflammasome activation via NEAT1 and MALAT1. Further evidence of inflammatory cell damage is seen in the downregulation of NUPR1 in BAL of M1 and M2 macrophages of severe patients with upregulation in mild patients. Downregulation of this stress response gene has been shown to cause mitochondrial dysfunction and ROS production that can lead to cell death.(70) Lastly, our observation that CTSL, a protein crucial for COVID-19 viral entry is upregulated across multiple cell types in severe patients provides a potential initial mechanism for the induction of the NEAT1 and MALAT1 mediated inflammatory state through increased efficiency of viral entry.(57)

Limitations in our study include the small sample size, variable clinical presentation and treatment. Additionally, time from presentation to sample collection varied across patients. The stratification of patients as severe or mild may also introduce unknown factors due to patient variability in presentation and classification. Although our validation shows promising reproduction of expression patterns, additional studies with more subjects and stringent recruiting and sample collection would further elucidate these findings.

We have demonstrated a clear ensemble of differential gene activity associated with severe disease in COVID-19 infection that revolves around the lncRNAs NEAT1 and MALAT1. Their specific activity changes in severe patients coupled with inflammasome promoting functions, suggest important roles in the COVID-19 hyperinflammatory process. These findings indicate that NEAT1 and MALAT1 may be candidates for treatment targeting or biological marker exploration.

## Data Availability

Data for BAL and PBMCs were downloaded from the NCBI Gene Expression Omnibus (accession number GSE145926) and the COVID-19 Cell Atlas (https:/www.covid19cellatlas.org/#wilk20), respectively. Data for validation was donwloaded from the following sources: nasopharyngeal validation - EGAS00001004481 (data downloaded from https://doi.org/10.6084/m9.figshare.12436517), and PBMC validation - EGAS00001004571(downloaded from https://beta.fastgenomics.org/datasets/detail-dataset-952687f71ef34322a850553c4a24e82e#Files)

https://www.ncbi.nlm.nih.gov/geo/query/acc.cgi?acc=GSE145926

## Supplementary Figures

**S1 Figure: Filtering of genes with extremely high residual variance. A**. A plot of mean expression levels versus residual variance for all genes detected in dataset. Genes with the highest residual variance are nearly all immunoglobulin and ribosome genes. **B**. Plot of 100 genes with highest residual variance in descending order. The rate of variance decrease stabilizes after the 21^st^ gene. C. Table of top 21 genes filtered out of downstream analysis due to very high residual variance.

**S2 Figure: Unbiased clustering of combined BAL and PBMC data reveals common cell types between cohorts and sample conditions. A/B**. UMAP plots showing the distribution of all cells in both cohorts within defined cell types. Plot A shows the 15 major cell groups we identified with labels over the cluster centers. Plot **B** shows the subclusters making up those groups. The major groups are abbreviated as prefixes with a letter suffix indicating subgroup. M1, M2 and Int are the macrophage/monocytes. NK is natural killer cells, E/G is epithelial cells and granulocytes, and E/P/C is epithelia, pneumocyte, and ciliary cells. **C**. Select markers for major cell groups plotted on the same UMAP projection. This illustrates the specificity of these markers for different regions of the plot corresponding to our respective cell types. D. Dot plot visualization of top markers utilized for identification of each cell cluster. The size of each dot indicates the percentage of cells with detectable expression of each gene, with color indicating expression level. **E**. Splitting the UMAP by cohort and by severity, with “H” indicating healthy control, “M” indicating mild disease, and “S” indicating severe disease illustrates that cell clusters do not organize according to sample type or patient condition, indicating successful integration of the datasets.

**S3 Figure: Transferred cell identities correspond to original cell identities from the nasopharyngeal validation set**. Dot plot shows cells from the nasopharyngeal validation dataset, identified by their transferred labels on the Y axis and their original labels on the X axis. Color of each dot indicates the numeric log frequency of cells that fit each corresponding set of labels. The size of each dot represents the percentage of each transferred cell types which is represented by each original cell type. From left to right, the full cell type names from the nasopharyngeal dataset are: non-resident macrophage (nrMa), monocyte-derived macrophage (MoD-Ma), monocyte-derived dendritic cell (moDC), resident macrophage (rMa), cytotoxic T cell (CTL), regulatory T cell (Treg), natural killer T cell (NKT), proliferating natural killer T cell (NKT-p), natural killer cells (NK), neutrophils (Neu), B cells, plasmacytoid dendritic cell (pDC), basal cell, ciliated cell, differentiating ciliated cell, FOXN4+ epithelial cell, ionocyte, interferon responsive cell (IRC), mast cell (MC), epithelial outliers, secretory, differentiating secretory, squamous, and unknown epithelial.

**S4 Figure: Transferred cell identities correspond to original cell identities from the PBMC validation set**. Dot plot shows cells from the PBMC validation set, identified by their transferred labels on the Y axis and their original labels on the X axis. Color of each dot indicates the numeric log frequency of cells that fit each corresponding set of labels. The size of each dot represents the percentage of each transferred cell types which is represented by each original cell type. This dataset contained several cell types where the same label was applied to more than one subcluster, resulting in numeric suffixes for similar cell types. mDCs correspond to myeloid dendritic cells and pDCs correspond to plasmacytoid dendritic cells.

## Supplementary Excel Tables

**S1 Table: Key characteristics of patients within each dataset**. Patient information from the BAL and PBMC cohorts used in this analysis. Patients who were intubated or had PaO2/FiO2 ⍰≤⍰300 mmHg were classified as severe. In BAL, patient “Mild 3” had only 369 cells recovered after filtering and was only used for initial clustering. Most patients in the BAL cohort. Exact ages were not available in the PBMC cohort. The first patient in this cohort was sampled twice, once while classified as a mild patient, and once after their symptoms worsened and required mechanical ventilation. Several patients in the PBMC cohort received azithromycin, which can have immunomodulatory effects before sample collection.

**S2 Table: Demographic characteristics of healthy subjects**. All healthy controls used from both the BAL and PBMC cohorts are listed.

**S3 Table: Cell identities from original datasets versus new combined identification**. For BAL and PBMCs, each cell’s original cell identification and new identification are tabulated for the 26 clusters in the subgroup sheets, and the 15 consolidated cell groups in the coarse sheets.

**S4 Table: P-value tables for cell proportion comparisons across each cell type**. Conditions compared are listed in the first two columns, and FDR adjusted p-values are listed in the third column. Each sheet is labeled by cell type.

**S5 and S6 Tables: DEGs for BAL and PBMC samples respectively, separated by cell type, with raw p-values as well as FDR adjusted p-values. Each sheet is labeled by cell type**. Column headings include indicators for which conditions are being compared where applicable. The conditions are numbered: “1=healthy control”, “2=mild COVID-19 patient”, “3=severe COVID-19 patient”. For example, the prefix “g2_1” indicates the comparison of mild patient expression levels minus healthy control expression levels. Log-fold change is reported relative to the natural log. Columns labeled “pct.1”, “pct.2”, or “pct.3” indicate the percentage of cells in the condition corresponding to that number with detectable expression of a particular gene.

**S7 Table: rDEGs with their expression levels in each cell type where each rDEG’s adjusted p-values passed the p-value filter as defined in our Methods section**. Column headings include indicators for which conditions are being compared where applicable. The conditions are numbered: “1=healthy control”, “2=mild COVID-19 patient”, “3=severe COVID-19 patient”. For example, the prefix “g2_1” indicates the comparison of mild patient expression levels minus healthy control expression levels. Log-fold change is reported relative to the natural log. Columns labeled “pct.1”, “pct.2”, or “pct.3” indicate the percentage of cells in the condition corresponding to that number with detectable expression of a particular gene. The “celltype” and “sample” columns indicate which cell type and which sample condition the rDEG passed filter in.

**S8 Table: Comparisons between differentially expressed rDEGs discussed in our results shows strong agreement in validation datasets**. A/B. Tables representing the tally of differential expression results of our discussed rDEGs which agreed or disagreed between analysis and validation groups based on the direction of detected differential expression. Tables are split by BAL/nasopharyngeal and PBMC groups. The first three columns correspond to cases where a comparison is not available due to a lack of differential expression detected in the original analysis (na.orig), the validation set (na.val), or both (na.all). The top half of each table reports the results for severe vs mild cases only (SvsM) while the bottom half reports results for all three comparisons: healthy vs mild, healthy vs severe, and severe vs mild.

**S9 and S10 Tables: Per gene level validation tables for BAL and PBMC groups respectively**. Each sheet name corresponds to the cell type presented. Row names indicate the gene being compared, and column names indicate the cohorts being compared: healthy (H), mild COVID (M), severe COVID (S). When differential expression is detected in both the original analysis and validation, the column is labled agree if the change occurred in the same direction and disagree if it is opposite. Other labels indicate where a comparison is not available due to a lack of differential expression detected in the original analysis (na.orig), the validation set (na.val), or both (na.all).

