## Supplementary figures and images for "Long non-coding RNAs (lncRNAs) NEAT1 and MALAT1 are differentially expressed in severe COVID-19 patients: An integrated single cell analysis"

### FigS1.bmp

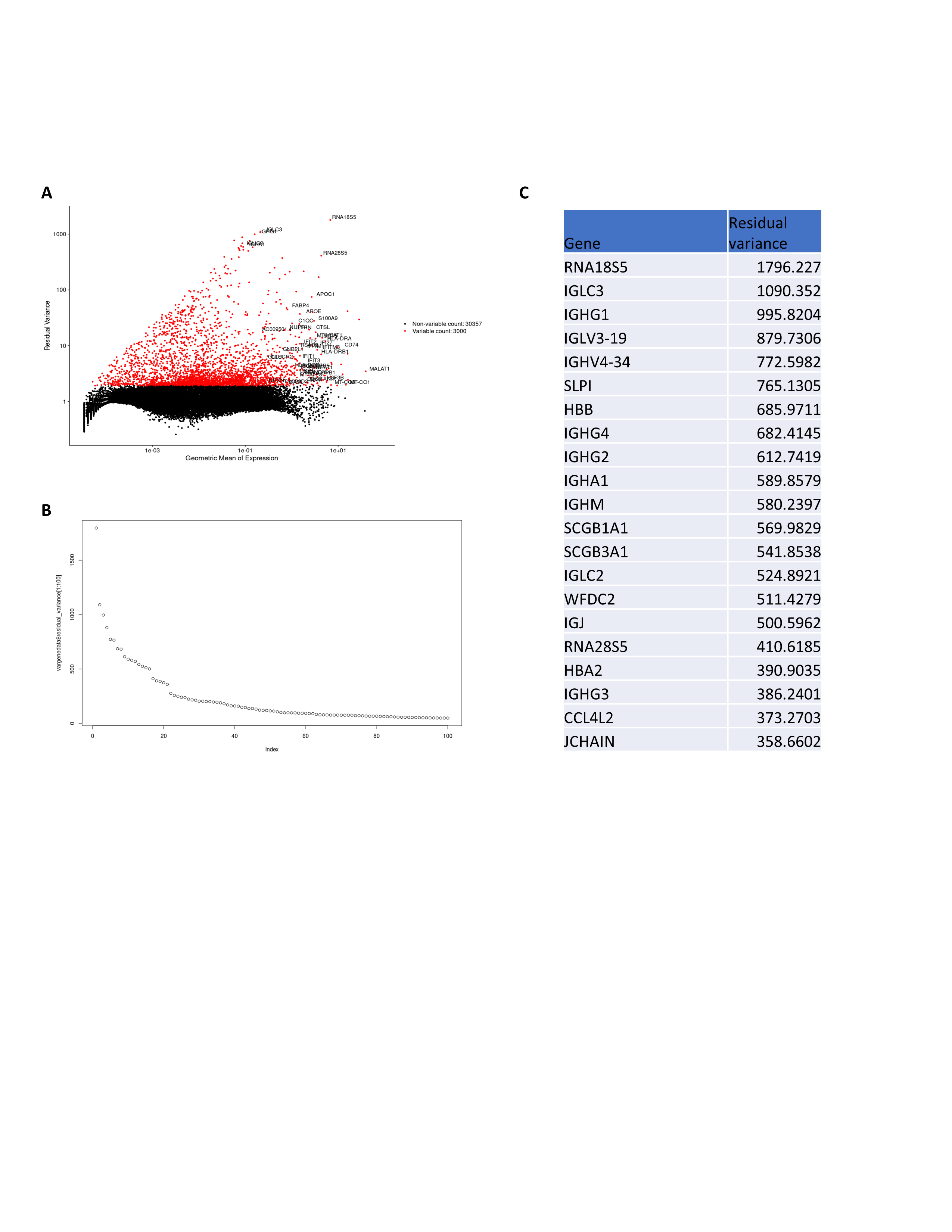

### FigS2.bmp

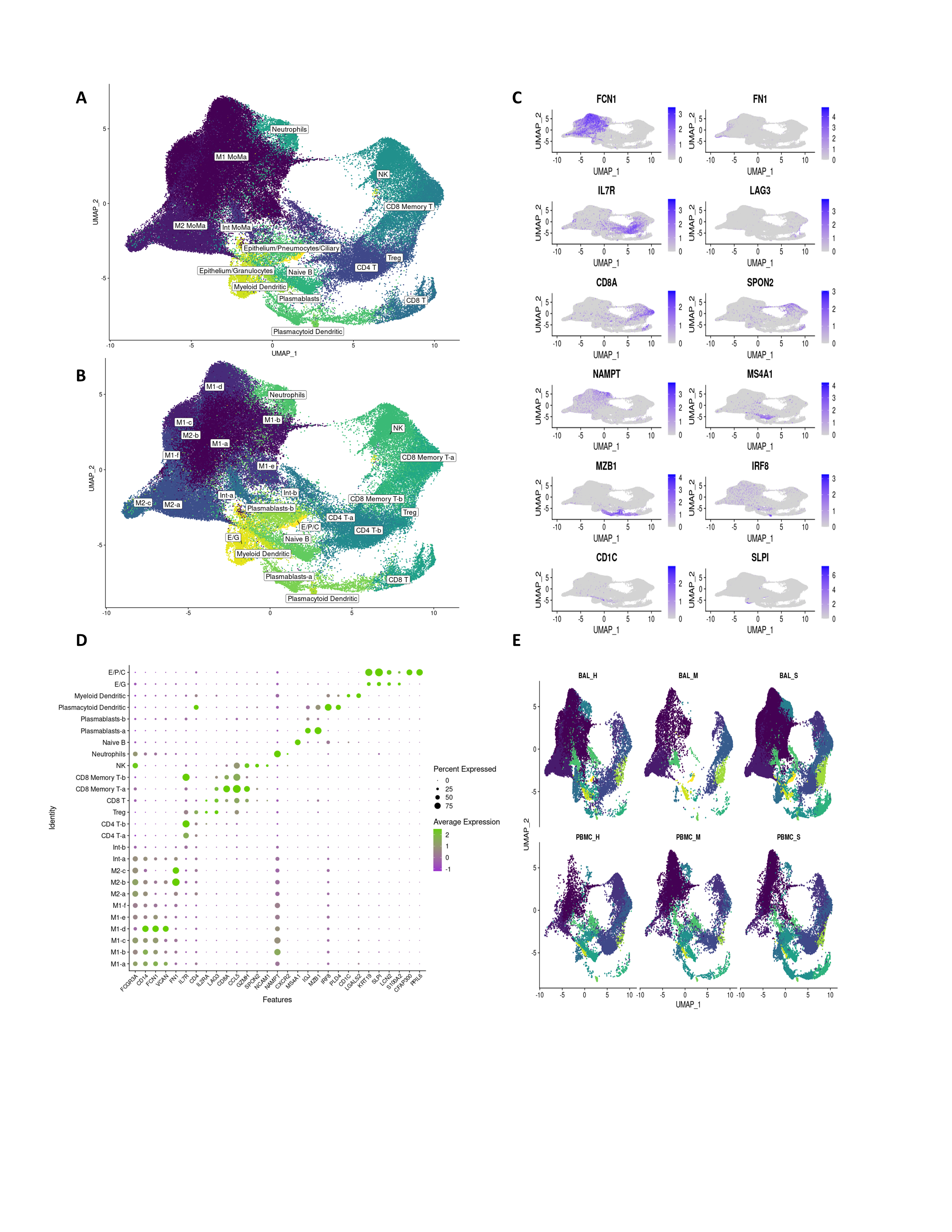

### FigS3.bmp

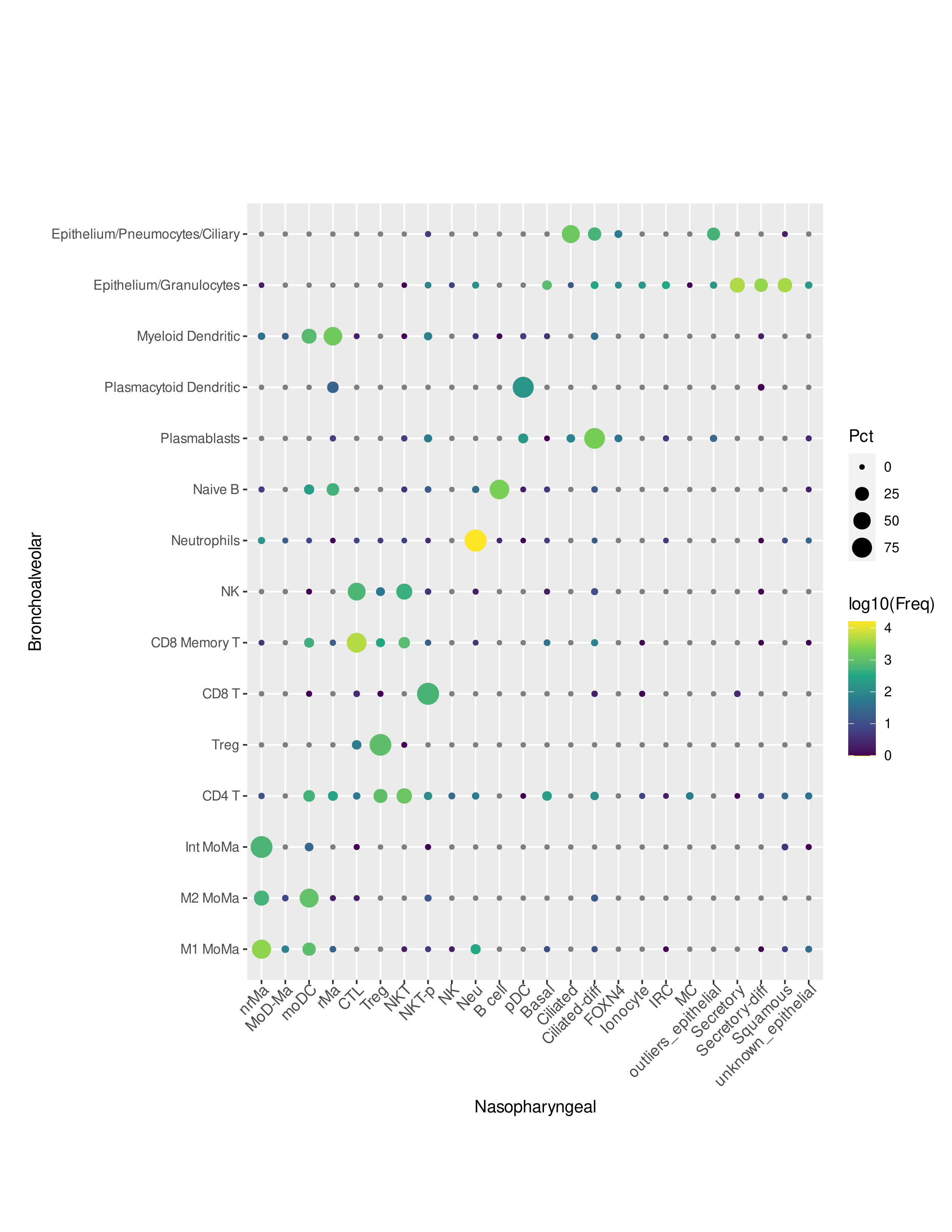

### FigS4.bmp

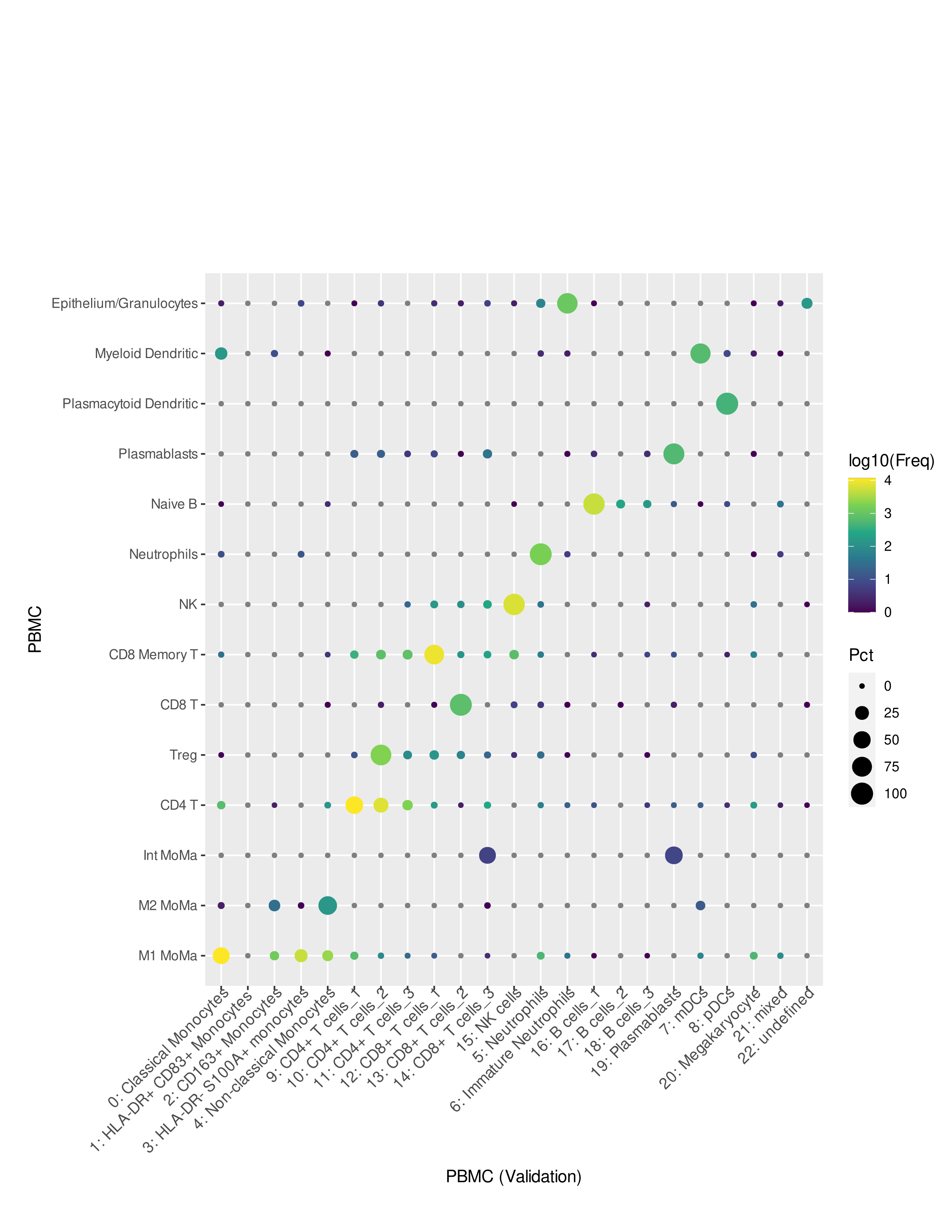
