## Supplemental Tables S1-S2 for "Long non-coding RNAs (lncRNAs) NEAT1 and MALAT1 are differentially expressed in severe COVID-19 patients: An integrated single cell analysis"

**Table S1: Key characteristics of patients within each dataset**. Patient information from the BAL and PBMC cohorts used in this analysis. Patients who were intubated or had PaO2/FiO2 ≤ 300 mmHg were classified as severe. In BAL, patient “Mild 3” had only 369 cells recovered after filtering and was only used for initial clustering. Most patients in the BAL cohort. Exact ages were not available in the PBMC cohort. The first patient in this cohort was sampled twice, once while classified as a mild patient, and once after their symptoms worsened and required mechanical ventilation. Several patients in the PBMC cohort received azithromycin, which can have immunomodulatory effects before sample collection.

| Subject | Cohort | Age | Gender | Sample Time (Days After Onset) | Interferon | Ribavirin | Methylprednisone | Azithromycin | Outcome |
|---|---|---|---|---|---|---|---|---|---|
| **Mild 1** | BAL | 36 | male | 11 | yes | yes | no | - | Discharged |
| **Mild 2** | BAL | 37 | female | 9 | no | no | no | - | Discharged |
| **Mild 3** | BAL | 35 | male | 13 | yes | yes | no | - | Discharged |
| **Severe 1** | BAL | 62 | male | 11 | yes | yes | no | - | Discharged |
| **Severe 2** | BAL | 66 | male | 18 | yes | yes | yes | - | Deceased |
| **Severe 3** | BAL | 63 | male | 14 | yes | yes | yes | - | Deceased |
| **Severe 4** | BAL | 65 | female | 15 | yes | yes | yes | - | Discharged |
| **Severe 5** | BAL | 57 | female | 8 | yes | no | yes | - | Discharged |
| **Severe 6** | BAL | 46 | male | 11 | yes | yes | no | - | Discharged |
| **Mild 1** | PBMC | 60-69 | male | 9 | - | - | - | yes | Discharged |
| **Severe 1** | | | | 11 | | | | | |
| **Mild 2** | PBMC | 40-49 | male | 16 | - | - | - | no | Discharged |
| **Mild 3** | PBMC | 50-59 | male | 15 | - | - | - | no | Discharged |
| **Mild 4** | PBMC | 20-29 | male | 12 | - | - | - | no | Discharged |
| **Severe 2** | PBMC | 30-39 | male | 9 | - | - | - | yes | Discharged |
| **Severe 3** | PBMC | 30-39 | male | 9 | - | - | - | yes | Discharged |
| **Severe 4** | PBMC | >80 | male | 2 | - | - | - | no | Deceased |

**Table S2: Demographic characteristics of healthy subjects.** All healthy controls used from both the BAL and PBMC cohorts are listed.

| Subject | Cohort | Age | Gender |
|---|---|---|---|
| **Healthy control 1** | BAL | 38 | female |
| **Healthy control 2** | BAL | 24 | male |
| **Healthy control 3** | BAL | 22 | male |
| **Healthy control 1** | PBMC | 49 | female |
| **Healthy control 2** | PBMC | 49 | male |
| **Healthy control 3** | PBMC | 36 | female |
| **Healthy control 4** | PBMC | 49 | male |
| **Healthy control 5** | PBMC | 48 | male |
| **Healthy control 6** | PBMC | 37 | male |
