## Supplemental Table S8 for "Long non-coding RNAs (lncRNAs) NEAT1 and MALAT1 are differentially expressed in severe COVID-19 patients: An integrated single cell analysis"

**A**

| BAL SvsM | na.orig | na.val | na.all | disagree | agree |
|---|---|---|---|---|---|
| M1 MoMa | 0 | 1 | 0 | 4 | 7 |
| M2 MoMa | 0 | 1 | 0 | 3 | 8 |
| NK | 0 | 9 | 1 | 1 | 1 |
| CD4 T | 0 | 4 | 0 | 2 | 6 |
| CD8 Memory T | 0 | 6 | 0 | 1 | 4 |
| **BAL All** | **na.orig** | **na.val** | **na.all** | **disagree** | **agree** |
| M1 MoMa | 0 | 22 | 0 | 4 | 10 |
| M2 MoMa | 0 | 25 | 0 | 3 | 8 |
| NK | 0 | 33 | 1 | 1 | 1 |
| CD4 T | 0 | 25 | 0 | 3 | 8 |
| CD8 Memory T | 0 | 27 | 1 | 1 | 4 |

**B**

| PBMC SvsM | na.orig | na.val | na.all | disagree | agree |
|---|---|---|---|---|---|
| M1 MoMa | 0 | 1 | 0 | 0 | 5 |
| M2 MoMa | 3 | 0 | 1 | 0 | 1 |
| NK | 0 | 1 | 0 | 0 | 4 |
| CD4 T | 0 | 1 | 0 | 1 | 2 |
| CD8 Memory T | 1 | 0 | 0 | 0 | 2 |
| **PBMC All** | **na.orig** | **na.val** | **na.all** | **disagree** | **agree** |
| M1 MoMa | 0 | 3 | 0 | 4 | 11 |
| M2 MoMa | 3 | 10 | 1 | 0 | 1 |
| NK | 0 | 3 | 0 | 1 | 11 |
| CD4 T | 0 | 3 | 1 | 1 | 7 |
| CD8 Memory T | 2 | 0 | 0 | 0 | 7 |
